# Heritability and rare variant contributions to Alzheimer disease in the Midwestern Amish

**DOI:** 10.64898/2026.09.17.26363309

**Authors:** Yining Liu, Yeunjoo E. Song, Weihuan Wang, Audrey Lynn, Kristy Miskimen, Sarada L. Fuzzell, Sherri D. Hochstetler, Renee A. Laux, Dawn Miller, Penelope Miron, Laura J. Caywood, Jason E. Clouse, Sharlene D. Herington, Ping Wang, Alex Gulyayev, Daniel A. Dorfsman, Noel C. Moore, Dana Z. Jian, Leighanne R. Main, Michael B. Prough, Andrew F. Zaman, Larry D. Adams, Patrice Whitehead, Paula Ogrocki, Alan J. Lerner, Jeffery M. Vance, Michael L. Cuccaro, William K. Scott, Margaret A. Pericak-Vance, Jonathan L. Haines

## Abstract

**Background:** The genetic architecture of Alzheimer disease remains incompletely understood. Founder populations such as the Amish offer a unique opportunity to identify additional genetic variation influencing the disease.

**Method:** Using extensive pedigree and genomic data from Midwestern Amish communities, we estimated both pedigree- and SNP- based heritability of AD. In addition, genome-wide association studies and gene-based rare variant tests were performed.

**Results:** Heritability estimates suggested that common variants explain much of genetic risk among *APOE-*ε4 carriers, while unexplained heritability remains in non-carriers in the Amish. GWAS confirmed *APOE-*ε4 as the strongest genetic determinant of AD in the Amish. Rare variant analyses identified *RIN3* and *PILRA* as significantly associated with risk of cognitive impairment; suggestive signals including *CYP24A1* were also identified.

**Discussion:** These findings highlight the contributions of common and rare variants to Alzheimer disease and suggest factors beyond common variants may contribute to disease susceptibility particularly among the *APOE*-ε4 non-carriers.

## 1 Introduction

Alzheimer Disease (AD), the most common form of dementia, affects millions of people worldwide and current treatment options only modestly slow progression in a subset of individuals.^1^ Previous analyses demonstrate that AD is highly heritable, indicating that genetic variation plays a substantial role in disease susceptibility.^2^ Genome-wide association studies (GWAS) have identified numerous common genetic variations, underscoring the polygenic nature of the disease.^3–5^ Among these loci, the *APOE*-ε4 allele consistently represents the strongest common genetic risk factor. However, it explains only a modest portion of the heritable component of AD, suggesting additional associated genetic variation remains to be discovered.^6–8^

Estimates of narrow-sense AD heritability from previous SNP-based and twin-based studies range widely from 3% to 79%, further suggesting the substantial unexplained genetic risk.^6,9–11^ However, interpretation of these estimates is complicated by several limitations. First, they were derived from diverse cohorts with varying sample sizes and study designs, limiting direct comparisons and generalizability.^12^ Second, many prior family-based estimates used a twin study design, which relies on the equal environment assumption.^12^ This assumption could potentially lead to overestimation and may also be less applicable to populations with complex family structures.^13^ Lastly, very few studies across any disease have examined both SNP- (h^2^_SNP_) and pedigree- (h^2ped^) based heritability using the same data.

Founder populations with extensive pedigree information can provide a valuable opportunity to estimate narrow-sense heritability more directly. The Midwestern Amish represents a unique founder population, characterized by large pedigrees, reduced genetic heterogeneity, and relatively homogenous environmental exposures.^14,15^ The availability of detailed and large pedigree and genetic information in this population enables the estimation of both h^2^_SNP_ and h^2^_ped_ within the same individuals, providing a more comprehensive view of the genetic contribution to AD risk.

Beyond common variant effects, rare variants (RVs) may be potential contributors to missing heritability.^16,17^ RVs tend to have larger effect sizes and can be more deleterious than common variants, but their low population frequencies often make them difficult to detect.^18–20^ Nevertheless, multiple studies have identified rare variants associated with both AD risk and protection. RVs in *PSEN1*, *PSEN2*, and *APP* are known drivers of early-onset familial AD, while RVs in genes such as *SORL1*, *TREM2*, and *ABCA7* are associated with increased late-onset AD risk.^21–26^ Additionally, other rare variants confer protection against cognitive decline, such as the A673T variant in.^27^ Identifying rare variants associated with either disease risk or protection may therefore provide crucial insights into AD pathophysiology and reveal potential therapeutic targets.

The Amish founder population provides a powerful setting for such discovery. The frequency of some rare and deleterious variants could be enriched through both genetic bottleneck and genetic drift, allowing them to be detected more easily.^28^ For example, Amish-enriched variants in *PGD* are associated with shortened telomere length; these variants are very rare in the general population.^29^ This framework is valuable not only for discovering variants that increase AD risk, but also for identifying protective variants against cognitive decline. Our previous studies detected loci on chromosomes 1 and 2 associated with cognitive preservation, a phenotype observed in cognitively intact older individuals with first degree relatives who were cognitively impaired.^30,31^ In addition, a locus on chromosome 17 (*SHISA6*) has been associated with delayed cognitive impairment in the Amish and was replicated in a collection of non-Amish families of European Ancestry.^32^ Together, these findings support further investigation of RVs that may increase or decrease the risk of dementia in the Amish.

In this study, we leveraged extensive pedigree and genetic data from the Amish families to investigate the genetic architecture of AD, and cognitive impairment more generally, through the associations of common and rare variants. We estimated h^2^_SNP_ and h^2^_ped_ of AD, and we conducted GWAS and gene-based rare variant association tests (RVATs) to detect variants associated with cognitive impairment and AD. By integrating analyses of both common and rare variants, our study aims to provide new insights into the genetic determinants of cognitive impairment and AD.

## 2 Methods

### 2.1 Genotyping chip, imputation, and quality control (QC)

Blood samples from 3,108 participants were genotyped using either Multi-Ethnic Genotyping Array (MEGA^ex^) or Global Screening Array (GSA) genotyping chips. Prior to imputation, samples with a genotyping rate < 95%, sex discrepancies, or relationship errors were excluded. Variants with call rate < 95%, Mendelian errors, or monomorphic sites were also removed. A total of 337,984 variants and 3,108 samples passed this initial QC. Genotype imputation was performed against TOPMed-r3 reference panel aligned to GRCh38.^33,34^ After repeating the quality control procedures, the imputed dataset used in this study contained 14,361,332 variants (including 5,807,927 common variants) across 3,106 samples.

### 2.2 Whole-genome sequencing (WGS) and QC

1,151 blood samples from the study participants were whole-genome sequenced via Illumina NovaSeq6000 platform. All WGS were pre-processed following the Alzheimer’s Disease Sequencing Project (ADSP) calling pipelines, and the reads were aligned to GRCh38.^35^ For QC, samples with sex mismatch, ID mismatch, relationship errors, and call rate < 99% were excluded. Monomorphic SNPs, SNPs with call rate < 99%, and SNPs deviating from Hardy-Weinberg Equilibrium (p < 1×10^−8^) were excluded. We additionally excluded samples with genome quality (GQ) < 20 and definition of coverage (DP) < 10. Only biallelic autosomal SNPs were included in this study, and the QC procedures were performed using KING 2.3.4 and PLINK 1.9.^36,37^ A total of 1,134 samples and 16,436,783 variants passed QC.

### 2.3 Study population, cognitive screening, and phenotype definition

The study participants were Amish individuals from central Ohio and northern Indiana ascertained under the Collaborative Amish Aging and Memory Project (CAAMP).^38^ Enrolled study participants underwent a full battery of neurocognitive assessments, including the Modified Mini-Mental State (3MS) exam, AD8 Dementia Screening Interview, Trail Making Test, Word List Memory Tasks, Verbal Fluency, Logical Memory, and Multilingual Naming Test at the time of blood draw.^39–42^ The cognitive status was further evaluated and adjudicated by a clinical adjudication board based on neurocognitive and physical exams and medical history.

Cognitive status was classified into five categories: cognitively unimpaired (CU), mild cognitive impairment (MCI), cognitively impaired but not AD (CINAD), AD, and unclear. CU individuals were considered as cognitively preserved because they remained cognitively unimpaired despite their older ages and having first degree relatives with dementia, and only CU participants who were at least 65 years old at the time of examination were included in these analyses. For some of the analyses, participants classified as MCI, CINAD, and AD were together considered as cognitively impaired (CI); for analyses specific to AD, only participants with a cognitive status of AD were included. 1,553 participants aged 51-102 years with valid clinically adjudicated cognitive status were genotyped (**Table 1**), and 917 participants aged 62-101 years also had whole-genome sequencing available (**Table 2**).

**Table 1.** Sample characteristics of participants included in the GWAS.

|  | **Overall**<br>(N = 1553) | **CU**<br>(N = 1001) | **CI**<br>(N=552) | **CI Subgroups** |  |  |
| --- | --- | --- | --- | --- | --- | --- |
|  |  |  |  | **MCI**<br>(N = 234) | **CINAD**<br>(N = 90) | **AD**<br>(N = 228) |
| **Participants, n (%)** | 1553 | 1001(64.46) | 552 (35.54) | 234 (42.39) | 90 (16.30) | 228 (41.30) |
| **Sex, n (%)** |  |  |  |  |  |  |
| Male | 615 (39.60) | 386 (38.56) | 229 (41.49) | 102 (43.59) | 46 (51.11) | 81 (35.53) |
| Female | 938 (60.40) | 615 (61.44) | 323 (58.51) | 132 (56.41) | 44 (48.89) | 147 (64.47) |
| **Age at Exam, years** |  |  |  |  |  |  |
| Mean (SD) | 81.23 (6.98) | 79.68 (6.81) | 84.03 (6.39) | 83.30 (5.67) | 83.23 (7.51) | 85.11 (6.48) |
| Range | 51-102 | 65-102 | 51 -102 | 60 - 97 | 51 - 97 | 66 - 102 |
| **APOE genotype, n (%)** |  |  |  |  |  |  |
| $\epsilon 2/\epsilon 2$ | 1 (0.06) | - | 1 (0.18) | 1 (0.43) | - | - |
| $\epsilon 2/\epsilon 3$ | 132 (8.50) | 100 (9.99) | 32 (5.80) | 12 (5.13) | 7 (7.78) | 13 (5.70) |
| $\epsilon 2/\epsilon 4$ | 29 (1.87) | 21 (2.10) | 8 (1.45) | 2 (0.85) | 3 (3.33) | 3 (1.32) |
| $\epsilon 3/\epsilon 3$ | 1030 (66.32) | 697(69.63) | 333 (60.33) | 166 (70.94) | 56 (62.22) | 111 (48.69) |
| $\epsilon 3/\epsilon 4$ | 330 (21.25) | 176 (17.58) | 154 (27.92) | 50 (21.37) | 22 (24.44) | 82 (35.96) |
| $\epsilon 4/\epsilon 4$ | 30 (1.93) | 6 (0.60) | 24 (4.35) | 3 (1.28) | 2 (2.22) | 19 (8.33) |
| N/A | 1 (0.06) | 1 (0.10) | - | - | - | - |

**Table 2.** Sample characteristics of participants included in the whole-genome sequencing analyses.

|  | **Overall**<br>(N = 917) | **CU**<br>(N = 518) | **CI**<br>(N=399) | **CI subgroups** |  |  |
| --- | --- | --- | --- | --- | --- | --- |
|  |  |  |  | **MCI**<br>(N = 161) | **CINAD**<br>(N = 58) | **AD**<br>(N = 180) |
| **Participants, n (%)** | 917 | 518 (56.49) | 399 (43.51) | 161 (40.35) | 58 (14.54) | 180 (45.11) |
| **Sex, n (%)** |  |  |  |  |  |  |
| Male | 372 (40.57) | 202 (39.00) | 170 (42.61) | 73 (45.34) | 32 (55.17) | 65 (36.11) |
| Female | 545 (59.43) | 316 (61.00) | 229 (57.39) | 88 (54.66) | 26 (44.83) | 115 (63.89) |
| **Age at Exam, years** |  |  |  |  |  |  |
| Mean (SD) | 83.55 (5.66) | 82.52 (5.54) | 84.88 (5.54) | 84.20 (4.77) | 84.33 (5.65) | 85.66 (6.04) |
| Range | 62 - 101 | 65 - 101 | 62 - 102 | 67 - 97 | 62 - 93 | 66 - 101 |
| **APOE genotype, n (%)** |  |  |  |  |  |  |
| $\epsilon 2/\epsilon 2$ | 1 (0.11) | - | 1 (0.25) | 1 (0.62) | - | - |
| $\epsilon 2/\epsilon 3$ | 89 (9.71) | 63 (12.16) | 26 (6.52) | 9 (5.59) | 5 (8.62) | 12 (6.67) |
| $\epsilon 2/\epsilon 4$ | 20 (2.18) | 12 (2.32) | 8 (2.01) | 2 (1.24) | 3 (5.17) | 3 (1.67) |
| $\epsilon 3/\epsilon 3$ | 590 (64.34) | 357 (68.92) | 233 (58.40) | 113 (70.19) | 33 (56.90) | 87 (48.33) |
| $\epsilon 3/\epsilon 4$ | 202 (22.03) | 85 (16.41) | 117 (29.32) | 34 (21.12) | 17 (29.31) | 66 (36.67) |
| $\epsilon 4/\epsilon 4$ | 15 (1.64) | 1 (0.19) | 14 (3.51) | 2 (1.24) | - | 12 (6.67) |

### 2.4 Heritability estimation

Pedigree-based heritability of AD was estimated using the ASSOC program implemented in the Statistical Analysis for Genetic Epidemiology (S.A.G.E) software.^43^ Because the full 14-generation Amish genealogy is computationally intensive for variance-component estimation, h*^2^*_ped_ was estimated from grandparental-family pedigrees defined by shared grandparental lineage. SNP-based heritability was estimated using the TetraHer tool for related samples within the LDAK software, based on inferred genetic relatedness up to the second degree.^44^ Both h*^2^*_ped_ and h*^2^*_SNP_ estimates accounted for sex, age of exam, and study center as covariates.

### 2.5 Rare variants selection and annotation

We defined rare variants as those with both a minor allele frequency (MAF) ≤ 1% in the non-Finnish European population from gnomAD v.4.0 and a minor allele count (MAC) ≤ 114 in the Amish cohort (MAF ≤ 5%). Using these thresholds enabled us to retain Amish-enriched variants while ensuring they remain rare in the general European population. This strategy also improves interpretability of identified associations and their applicability to the general population.

Among 5,718,279 RVs observed in both gnomAD and the Amish dataset, 63,363 had MAC > 114 in the Amish and were analyzed as common variants. The remaining 5,654,916 RVs were included in downstream rare variant analysis, including 1,027,135 variants enriched in the Amish cohort, with Amish MAF between 1% and 5%. RVs were annotated by both SnpEff and FAVOR annotator to determine their putative functional impact.^45,46^

We selected RVs annotated by SnpEff as having high impact (e.g., predicted loss-of-function, pLOF) or moderate impact (e.g. missense variants) on protein function to test those affecting protein-coding abilities.^49^ High-impact pLoF variants are assumed to have disruptive impact on protein structure or function, while moderate-impact variants might alter protein effectiveness but are generally less severe than high-impact variants.

The FAVOR annotator was used to further classify disruptive missense variants and to annotate non-coding rare variants.^50^ RVs annotated as upstream, downstream, untranslated regions (UTR), promoters or enhancers, overlaid with DNase Hypersensitivity (DHS) sites and Cap Analysis of Gene Expression (CAGE) by the FAVOR annotator, were included for non-coding rare variant analyses following the STAARpipeline.^47,48^

### 2.6 Statistical analysis

For common variant association tests, we performed linear mixed models using the GENESIS Bioconductor package, accounting for sex, age of exam, study center, and a genetic relationship matrix (GRM).^49^ Genetic variants were considered genome-wide significant with p-values < 5 × 10^−8^ and genome-wide suggestive with p-values <1 × 10^−5^. Quantile-Quantile (QQ) plots were created using qqman R package.^50^

We tested both coding and non-coding variants for rare variant association tests. Coding RVs annotated as high-impact pLOF or disruptive missense variants were aggregated based on genes for burden tests. We used SAIGE-GENE+ for performing burden tests, optimal sequenced kernel association tests (SKAT-O), and single variant association tests.^51^ Variants with MAC < 5 were considered as ultra-rare (URV) and were collapsed together in this RVAT analysis. We also performed sensitivity analyses by expanding the RV list to include all high- and moderate-impact variants (SKAT-O test).

We followed the STAARpipeline for detecting RVs specifically in the non-coding regions of protein-coding genes.^47,48^ RVs in each of the seven functional categories were aggregated and tested. We used the STAAR-O p-values output correcting for the 7 masks when defining the significant and suggestive thresholds in this analysis.

For the RVATs, we also adjusted a set of covariates including sex, age of exam, center of sample collection, and GRM in all statistical analyses (model 1). We additionally adjusted for the presence of *APOE*-ε4 (model 2), and both the presence of *APOE*-ε4 and *APOE*-ε2 (model 3).

Only genes with at least two RVs of interest and with a cumulative MAC (cMAC) > 5 were examined. We set our significant threshold as 0.05/number of genes being tested, and suggestive thresholds as 1/number of genes being tested for each statistical test in each analysis.

All analyses were performed using two phenotypic contrasts: 1) CI vs CU to evaluate genetic associations with cognitive preservation and impairment of cognitive function; and 2) CU vs AD to evaluate genetic associations with AD risk. Due to phenotype coding, effect directions differ between two constructs: for CI vs CU, positive β indicates cognitive preservation and reduced risk, whereas for CU vs AD, positive β indicates increased AD risk. Although the CI vs CU construct was coded with CU as the outcome, the associations identified in this study were mainly in the direction of increased risk of cognitive impairment and were therefore described accordingly.

## 3 Results

### 3.1 Heritability of AD reveals missing heritability in *APOE*-ε4 non-carriers

To evaluate the contribution of common variants and to get an overview of the genetic architecture of AD in the Midwestern Amish, we integrated the extensive Amish pedigree structure and genetic information to estimate both pedigree-based and SNP-based heritability of AD (**Table 3**).

**Table 3.**
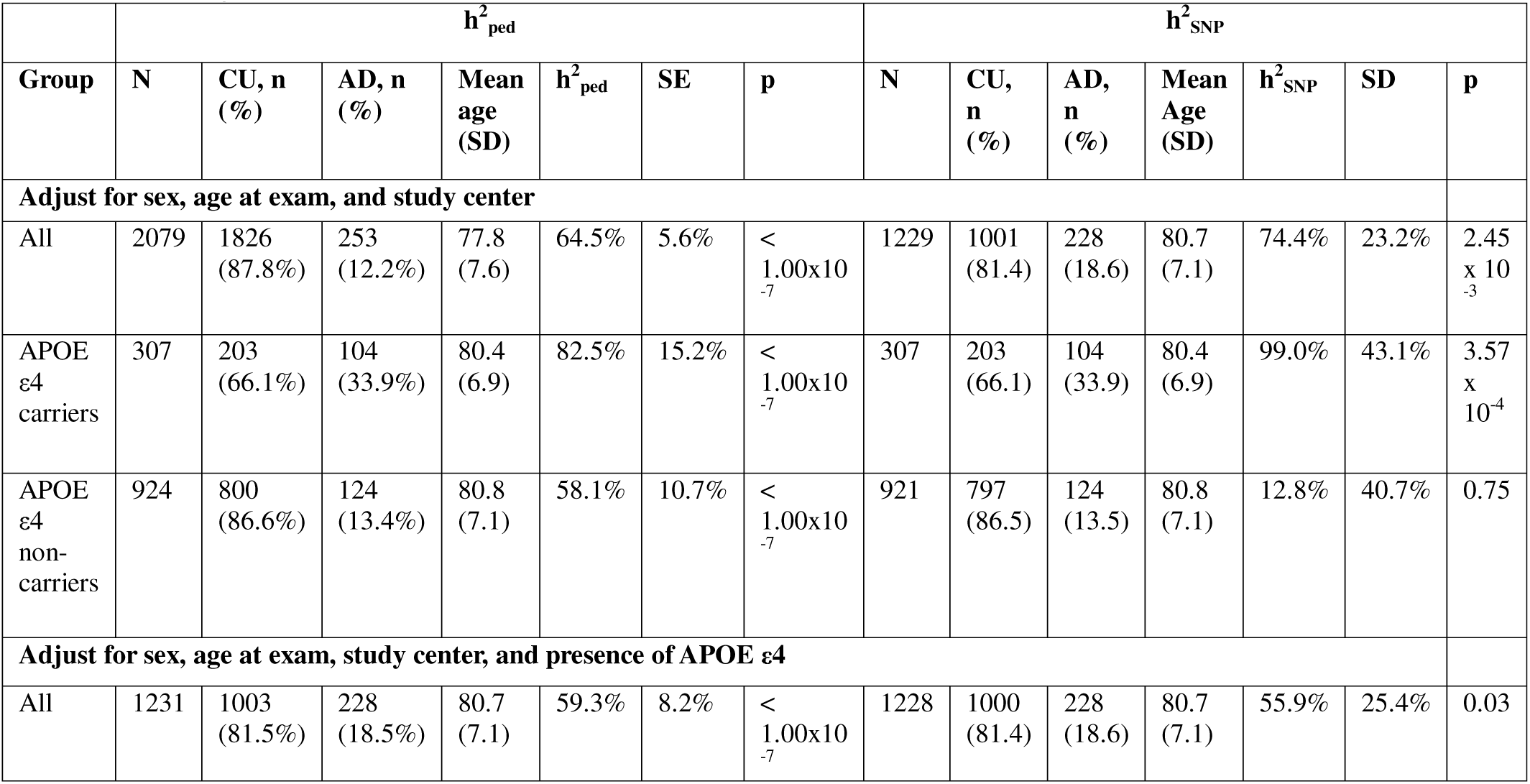
Pedigree- and SNP- based heritability of Alzheimer disease.

The pedigree-based heritability estimate for AD was 64.5% (SE = 5.6 %, p < 1.00 × 10^−7^), which decreased to 59.3% (SE = 8.2%, p < 1.00 × 10^−7^) after accounting for the presence of the *APOE*-ε4 allele. Stratified analyses showed that h^2^_ped_ was 82.5% (SE = 15.2%, p < 1.00 × 10^−7^) among *APOE*-ε4 carriers and 58.1% (SE = 10.7%, p < 1.00 ×10^−7^) among non-carriers.

The SNP-based heritability estimate for AD was 74.4% (SD = 23.2%, p =2.45 × 10^−3^), decreasing to 55.9% (SD = 25.4%, p =0.03) after accounting for the presence of the *APOE*-ε4 allele. Among *APOE*-ε4 carriers, h^2^ was estimated at 99.0% (SD = 43.1%, p = 3.57 × 10^−4^), whereas h^2^_SNP_ among non-carriers was 12.8% (SD = 40.7%, p = 0.75).

### 3.2 Association Analyses

#### 3.2.1 Common variant genome-wide association testing

We previously reported a GWAS of cognitive preservation in this population.^30^ Here we updated the GWAS analyses with additional participants and updated phenotype and genotype data. Sample characteristics for the GWAS are described in **Table 1**.

In the GWAS comparing 552 CI and 1,001 CU individuals, the strongest association signal was observed at rs429358 (chr19:44908648:T<C, p = 2.30 × 10^−12^), a defining variant of the *APOE*-ε4 allele (**Figure 1A**). Additional genome-wide significant signals in linkage disequilibrium with *APOE* were detected, including *TOMM40* and *NECTIN2* on chromosome 19 (p < 5 × 10^−8^), with all significant loci associated with increased risk of cognitive impairment (**Table S1**). No evidence of genomic inflation was observed (λ = 1.002, **Figure 1B**). Similarly, in the AD GWAS (1,001 CU and 228 AD individuals), genome-wide significant associations with increased risk were again concentrated in the *APOE* region, including *NECTIN2*, *TOMM40*, *APOC1*, *QPCTL*, and *HIF3A* (**Figure 1C**). The most significant signal was again observed at rs429358 (p = 1.02 × 10^−18^). Additional significant variants are summarized in **Table S2**. The QQ plot indicated minimal genomic inflation (λ = 1.016, **Figure 1D**).

**Figure 1.**
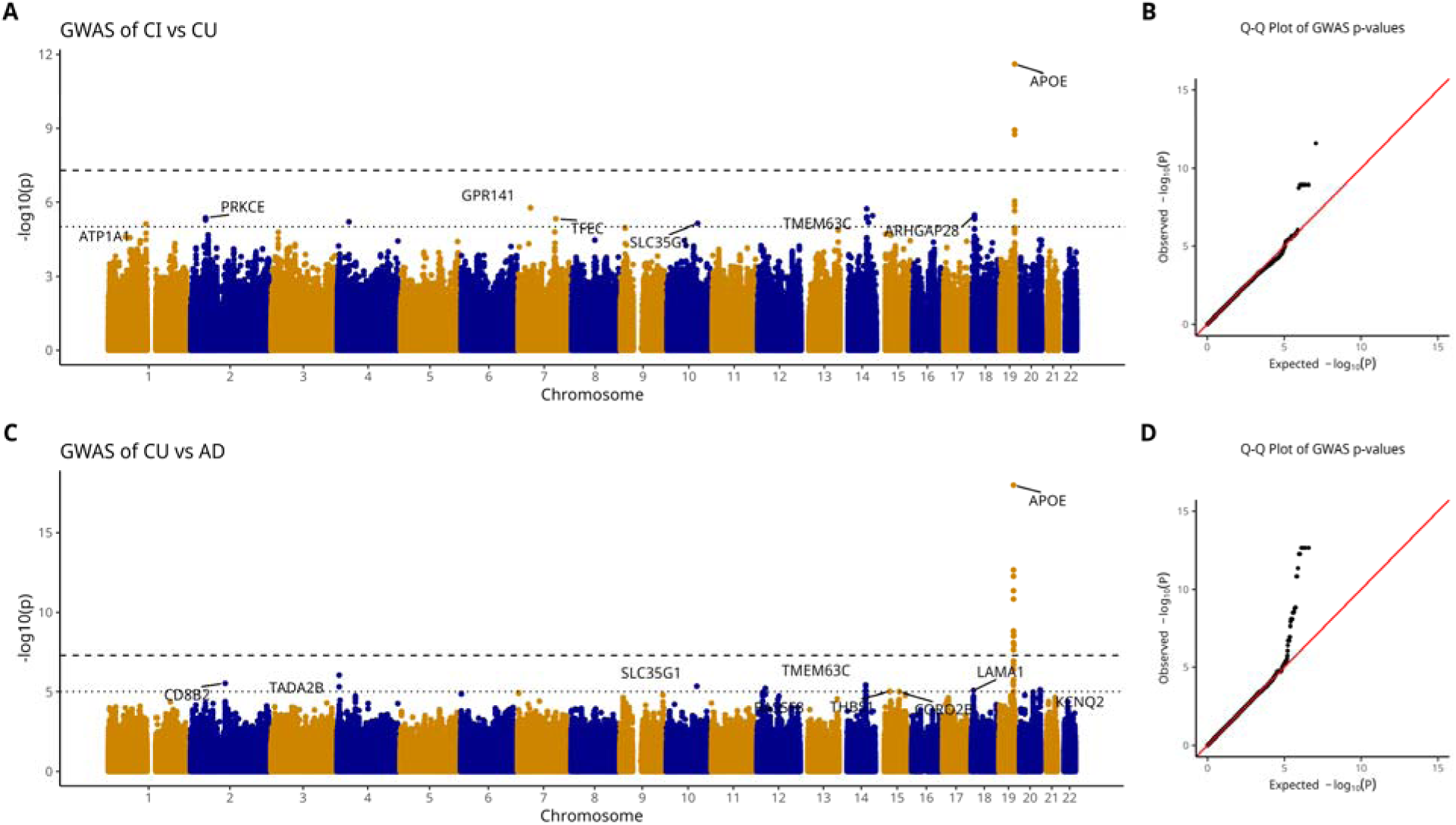
Genome-wide association analyses of cognitive preservation and AD. **A)** Manhattan plot showing the result of cognitive preservation GWAS. **B)** Q-Q plot of GWAS of cognitive preservation. λ = 1.002. **C)** Manhattan plot showing the result of AD GWAS. **D)** Q-Q plot of AD GWAS. λ = 1.016.

Stratified AD GWAS by *APOE*-ε4 carrier status revealed no genome-wide significant associations (**Figure S1 & S2**), likely due to reduced sample sizes. Nonetheless, among *APOE*-ε4 non-carriers, *VSTM2A* (rs10264395, p = 4.08 × 10^−7^) and *LRPAP1*(rs11549512, p = 2.83 × 10^−7^) showed the strongest suggestive associations, whereas signals observed in *APOE*-ε4 carriers were generally weaker and below the suggestive significance threshold.

#### 3.2.2 Rare variant association tests with known AD genes

Next, we performed rare variant association analyses using variants observed in previously defined AD-related genes to identify those genes in which rare variants may be associated with cognitive status in the Amish population. To reduce multiple testing burden and increase statistical power, we first prioritized our analyses to 20 known AD-related genes identified by the ADSP Gene Verification Committee (https://adsp.niagads.org/gvc-top-hits-list/). Sample characteristics for RVATs were summarized in **Table 2**.

Following variant annotation and filtering, only one of the 20 genes (*ABCA7*) met inclusion criteria for the primary analysis aggregating pLOF and disruptive missense rare variants.

However, variants in *ABCA7* showed no significant association in either CI vs CU (burden p = 0.91) or CU vs AD (burden p = 0.94). We therefore expanded the variant sets to include all RVs with high or moderate predicted impact, resulting in 7 genes tested in CI vs CU (**Table 4**) and 3 in CU vs AD (**Table S3**).

**Table 4.**
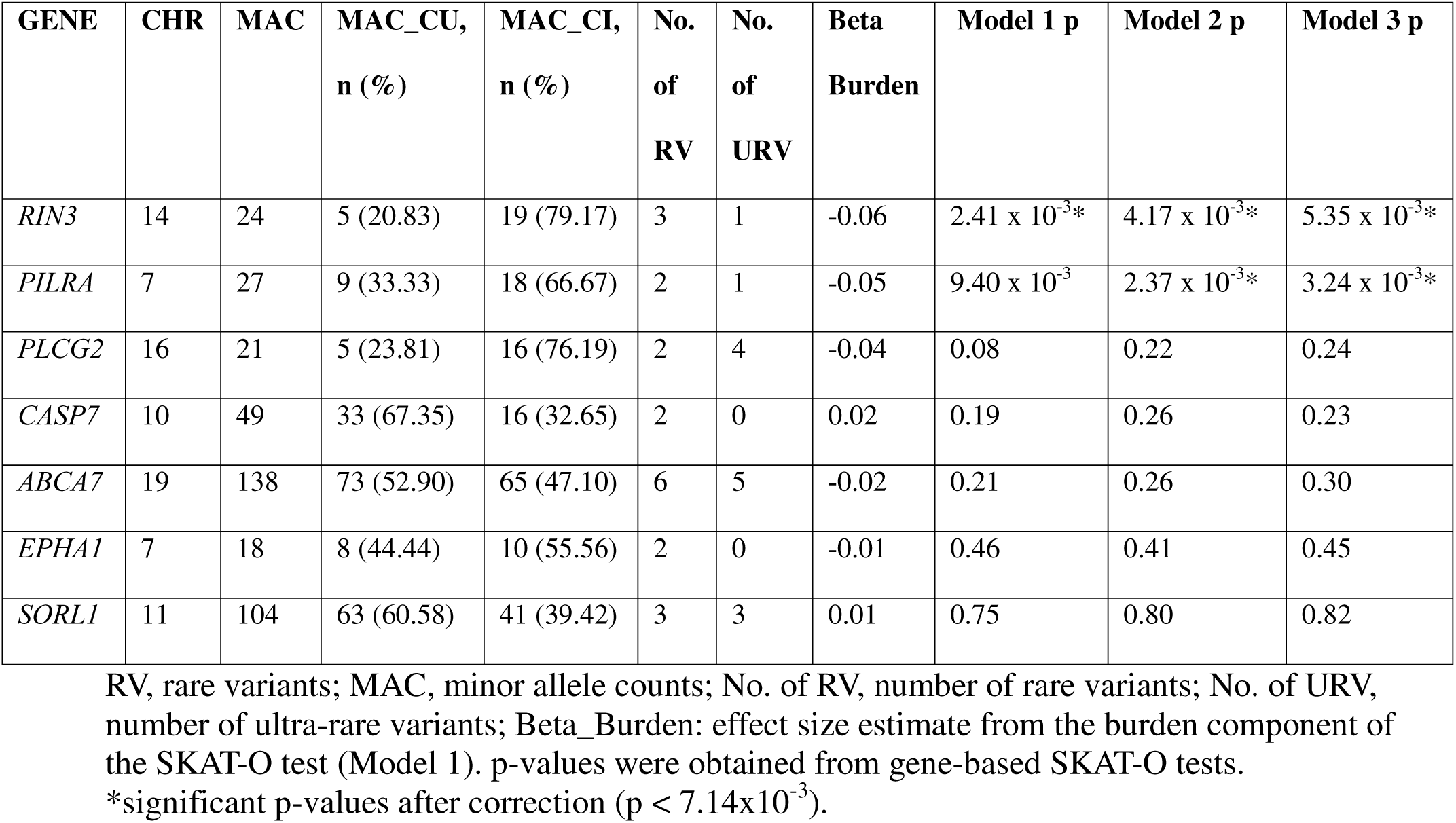
Gene-based rare variant association tests for CI vs CU in AD-related genes.

In the CI vs CU analysis, *RIN3* was significantly associated with increased risk of cognitive impairment across all models (model 1: SKAT-O p = 2.41 × 10^−3^, model 2: SKAT-O p = 4.17 × 10^−3^, model 3: SKAT-O p = 5.35 × 10^−3^), exceeding the Bonferroni-corrected significance threshold (0.05/7 = 7.14 × 10^−3^). *PILRA* also showed significant associations with risk in model 2 (p = 2.37 × 10^−3^) and model 3 (p = 3.24 × 10^−3^) after Bonferroni correction (**Table 4**). However, no significant associations were observed in the CU vs AD analysis (*ABCA7*: SKAT-O p = 0.09, *CASP7*: SKAT-O p =0.63, *SORL1*: SKAT-O p = 0.99, **Table S3**).

The four rare missense variants tested in *RIN3* were rs150221413 (chr14:92555895:G>T), rs149740709 (chr14:92652218:G>A), rs12434929 (ch14:92652887:G>C), and rs147042536 (chr14:92676516:T>C). In *PILRA*, three variants were tested: rs376057079 (chr7:100373659:G>C), rs201649203 (chr7:100397913:G>C), and rs201973358 (chr7:100399797:G>A), corresponding to an initiator codon variant, a splice donor variant, and a missense variant, respectively. Single-variant association tests were performed for non-ultra-rare variants (**Table S4**), in which rs150221413 in *RIN3* had a p-value of 2.65 × 10^−4^ (β = -3.98) and rs201973358 in *PILRA* had a p-value of 0.03 (β = -1.20).

#### 3.2.3 Genome-wide, gene-based rare variant association tests

We next performed genome-wide, gene-based burden tests by aggregating pLOF and disruptive missense variants within genes across autosomes.

In the CI vs CU analysis, *CYP24A1* showed suggestive association with increased risk of cognitive impairment both before and after adjusting for the effect of *APOE* (model 1: burden test p = 0.0024; model 2: burden test p = 0.0010, model 3: burden test p = 0.0015, **Figure 2A & B**). A consistent suggestive association was also observed in the CU vs AD analysis (model 1: burden p = 0.0038, model 2: burden p = 0.0014, model 3: burden p = 0.0016, **Figure 3A & B**). To examine whether this signal was driven by AD, we ran an additional burden test comparing CU and non-AD CI individuals, and a nominally significant burden p value of 0.018 was observed.

**Figure 2.**
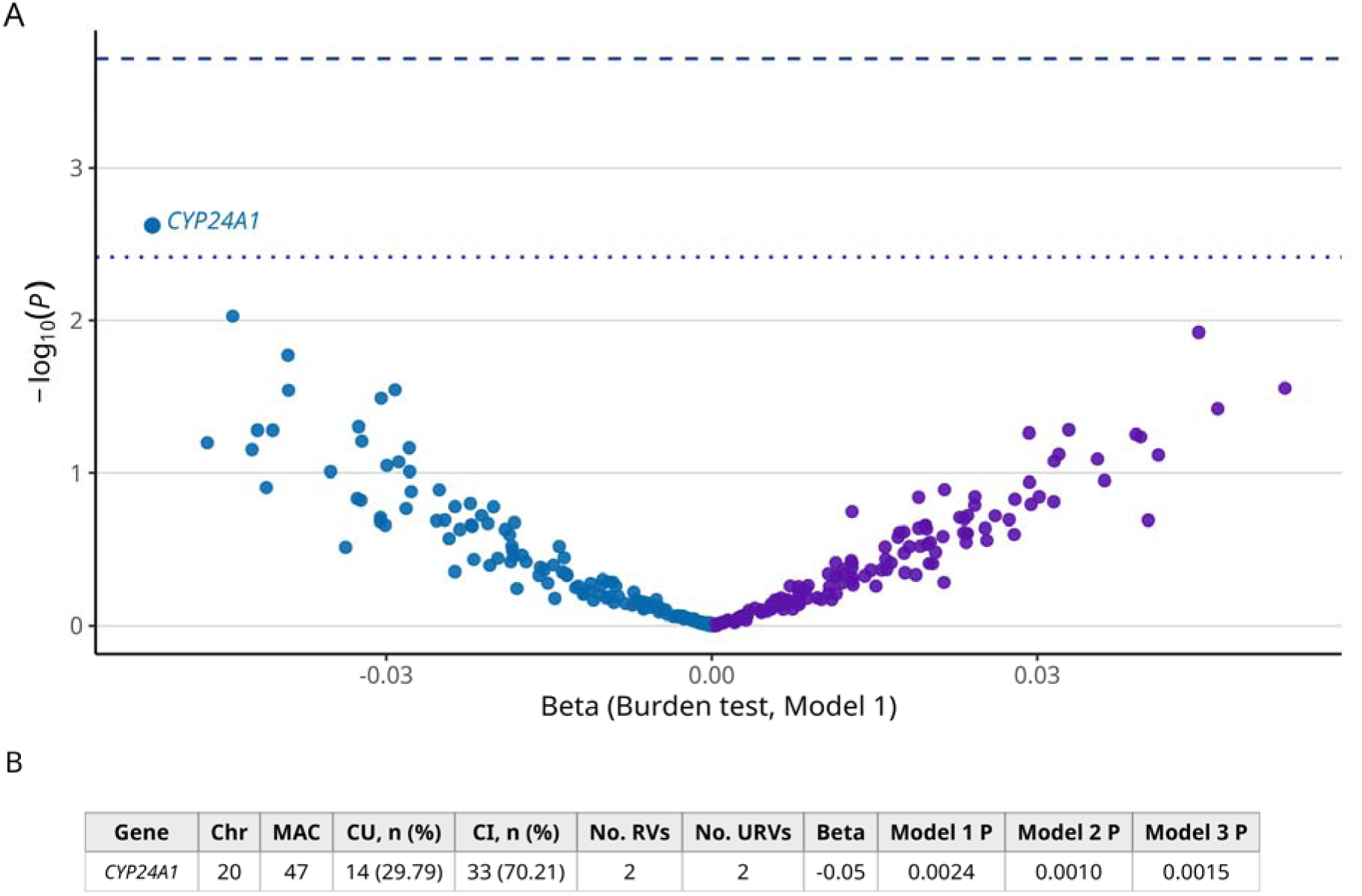
Gene-based rare variant association analysis of coding variants for CI vs CU. (**A) Volcano plot of gene-based burden tests for pLOF and disruptive missense RVs comparing CI and CU individuals**. Dashed line: genome-wide significance (−log□□(1.92 × 10□□)); dotted line: suggestive threshold (−log□□(0.0038)). Negative beta indicates increased risk of cognitive impairment. **(B) Detailed association results for *CYP24A1*.** MAC, minor allele counts; No. of RV, number of rare variants; No. of URV, number of ultra-rare variants; Beta_Burden: effect size estimate from the burden test (Model 1). A total of 260 genes were tested.

**Figure 3.**
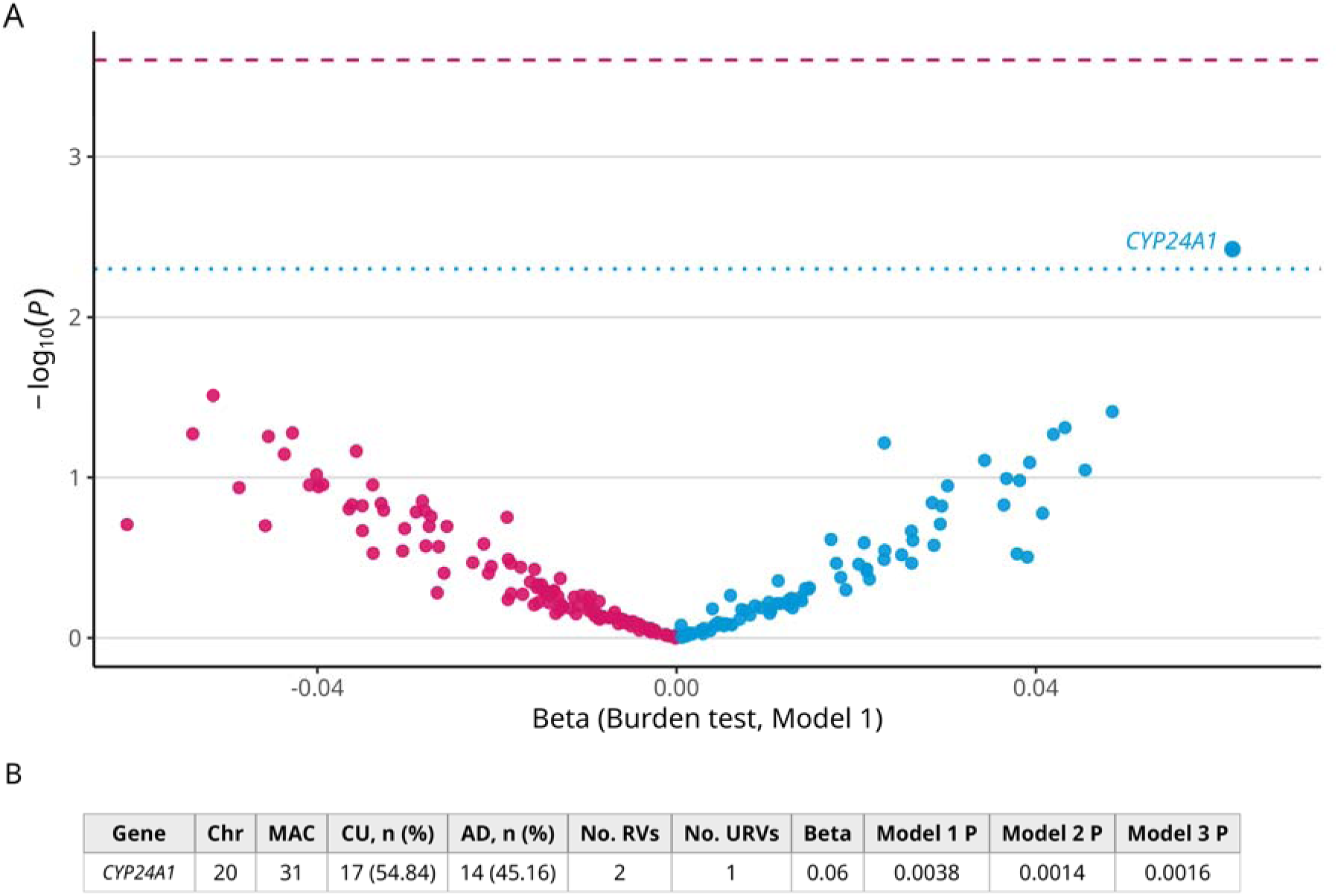
Gene-based rare variant association analysis of coding variants for CU vs AD. (**A) Volcano plot of gene-based burden tests for pLOF and disruptive missense RVs comparing CU and AD individuals**. Dashed line: genome-wide significance (−log□□(2.50 × 10^−4^)); dotted line: suggestive threshold (−log□□(0.0050)). Positive beta indicates increased AD risk. (B) Detailed association results for *CYP24A1*. MAC, minor allele counts; No. of RV, number of rare variants; No. of URV, number of ultra-rare variants; Beta_Burden: effect size estimate from the burden test (Model 1). A total of 200 genes were tested.

Furthermore, given prior evidence of unexplained heritability among *APOE*-ε4 non-carriers, we repeated the genome-wide burden tests in this subgroup. A total of 139 genes were tested, resulting in a Bonferroni-corrected significance threshold of 0.05/139= 3.60 × 10^−4^ and a suggestive threshold of 1/140 = 0.0072. Four loci reached suggestive significance, including *ACADS* (p = 0.0043), *CYP24A1* (p =0.0056), and *SLC6A18* (p =0.0069), all associated with increased risk, and *CP* (p =0.0066), which showed a protective direction (**Figure S3**).

Finally, gene-based RVATs of noncoding variants using the STAARpipeline identified several suggestive loci in both the CI vs CU and CU vs AD analyses, including *KLHDC1*, *TRAF3IP1*, *ACER2*, *LINC0092*, *CD3EAP*, *PPP1R13L*, and *ELOC* (**Table S5 & S6**).

## 4 Discussion

Leveraging the extensive pedigree structure and genetic data from the Amish families, we investigated the contributions of both common and rare variants to cognitive impairment and AD. Genome-wide association analyses confirmed *APOE*-ε4 as the primary risk locus for cognitive impairment in the Amish. Heritability estimates of AD indicated the presence of unexplained missing heritability for AD risk among *APOE*-ε4 non-carriers, and rare variant analyses further identified several loci that may contribute to disease risk in both all subjects and specifically among the *APOE*-ε4 non-carriers.

In the heritability analyses, pedigree-based heritability for AD was estimated to be approximately 64%, while SNP-based heritability was about 74%. Both pedigree- and SNP-based heritability estimates decreased after adjusting for the presence of *APOE*-ε4, consistent with previous estimates of AD heritability.^7,8,12^ The slightly higher, but non-significantly higher, SNP-based heritability compared to pedigree-based heritability among all subjects may likely be driven by the unique genetic architecture of the Amish founder population, including extended linkage disequilibrium and enrichment of shared variants.

Among *APOE*-ε4 carriers, h^2^_SNP_ was estimated as 99%, consistent with the known strong effect of *APOE*-ε4. h^2^_ped_ was also high in *APOE*-ε4 carriers, at about 82%. In contrast, h^2^_SNP_ was only about 13% and not statistically significant in the *APOE*-ε4 non-carriers, indicating that this group harbors much of the unexplained heritability for AD in the Amish, while h^2^_ped_ remained relatively high and statistically significant at 58% in the *APOE*-ε4 non-carriers. Because h^2^_SNP_ estimates were associated with relatively large standard deviations, the apparent differences should be interpreted with caution. Nevertheless, this discrepancy between h^2^_SNP_ and h^2^_ped_ suggests the presence of additional genetic and non-genetic factors among *APOE*-ε4 non-carriers. These may include genetic factors such as rare and structural variants, as well as non-genetic influences including lifestyle, environmental exposure, and metabolic conditions, together with gene-environment interactions.^52,53^

Analysis of rare variants focused on the known AD-related genes identified *RIN3* and *PILRA* as significantly associated with the risk of cognitive impairment in the Amish. Both genes have previously been implicated in AD risk and protection through common variant association studies.^3,54–58^ Single-variant tests suggested that the missense variant rs150221413 may drive the signal observed in *RIN3*. This missense variant was previously suggestively associated with both early- and late-onset AD risk.^59,60^ These observations suggest that this missense variant may act as a potential risk modifier.

For *PILRA*, there was not a single rare variant that clearly drove the association signal. Patel et al (2019) previously reported a significant burden of five rare variants in *PILRA* associated with AD.^61^ However, the three RVs tested in this study did not overlap with those reported in that analysis. These findings nevertheless support the possibility that rare variants in *PILRA* may contribute to the genetic architecture of cognitive impairment in addition to the effects of common variants.

In addition to known AD genes, rare variant burden tests showed a suggestive association in *CYP24A1* with cognitive impairment. The lack of statistically significant findings is likely due to limited sample sizes. Notably, this association was not specific to AD based on comparisons between CU and non-AD impaired individuals, suggesting that rare variants in this gene may influence the risk of broader cognitive impairment. *CYP24A1* encodes mitochondrial enzyme 24-hydroxylase, which plays an important role in the inactivation of active vitamin D3 and its precursor.^62^ Although the role of vitamin D in Alzheimer disease is unclear, several previous studies have reported that low vitamin D levels are associated with an increased risk of AD.^63–66^ Because vitamin D levels are strongly influenced by ultraviolet exposure from sunlight, the Amish lifestyle, characterized by substantial outdoor activity and agricultural work, may result in greater sunlight exposure, which in turn may interact with the genetic variation in *CYP24A1* to influence circulating vitamin D levels and related phenotypes, including cognition.^67^ Therefore, these observations suggest that the relationship between *CYP24A1*, vitamin D metabolism, and cognition warrants further investigation.

Although we detected only suggestive common and rare variant associations particularly among *APOE*-ε4 non-carriers possibly due to limited power, the top genes identified in these analyses have previously been implicated in dementia and other neurological conditions. *LRPAP1* and *VSTM2A*, identified in GWAS, were both suggestively associated with AD.^68–71^ *LRPAP1* regulates an apolipoprotein E receptor involved in amyloid-β clearance, whereas *VSTM2A*, despite having no established role in AD, may be involved in blood-brain-barrier regulaton.^68,72^ The *CP* gene, identified in RVAT as potentially protective against AD, encodes ceruloplasmin which is a key enzyme that regulates copper and iron metabolism. While the role of iron homeostasis in neurodegenerative diseases remains unresolved, accumulating evidence suggests the metal metabolism may contribute to neurological functions, and our findings stress the need for and importance of further investigation.^73–75^

Overall, our results suggest that the *APOE* region on chromosome 19 remains the most prominent locus associated with AD risk, while rare variants in known AD genes may also contribute somewhat to disease risk in the Amish. Despite the limitations, our study identified several loci that may contribute to cognitive decline, highlighting potential targets for future validation. Additional genetic and non-genetic contributions to AD risk likely remain to be discovered, particularly among *APOE*-ε4 non-carriers. Importantly, our findings underscore the value of studying rare variants in founder populations such as the Amish and emphasize the importance of investigating both risk and protective variants to fully understand the biological underpinnings of AD.

## Supporting information

All Supplementary

## Data Availability

All data produced in the present study are available upon reasonable request to the authors

## Acknowledgement

The authors would like to thank all the Amish families who participated in our study. The authors would like to extend our gratitude to Dr. Xihao Li from University of North Carolina – Chapel Hill for the guidance of running STAARpipeline. The authors would like to also acknowledge the support from the High-Performance Computing Resources team at Case Western Reserve University.

## Consent Statement

All study procedures were approved by the Institutional Review Board (IRB) at the Case Western Reserve University and University of Miami, and all individuals enrolled in this study supplied informed consent.

## References

1. Dyck CHv, Swanson CJ, Aisen P, et al. Lecanemab in Early Alzheimer’s Disease. New England Journal of Medicine. 2023;388(1):9–21. doi:doi:10.1056/NEJMoa2212948

2. Andrews SJ, Renton AE, Fulton-Howard B, Podlesny-Drabiniok A, Marcora E, Goate AM. The complex genetic architecture of Alzheimer’s disease: novel insights and future directions. eBioMedicine. 2023;90doi:10.1016/j.ebiom.2023.104511

3. Bellenguez C, Küçükali F, Jansen IE, et al. New insights into the genetic etiology of Alzheimer’s disease and related dementias. Nature Genetics. 2022/04/01 2022;54(4):412-436. doi:10.1038/s41588-022-01024-z

4. Kunkle BW, Grenier-Boley B, Sims R, et al. Genetic meta-analysis of diagnosed Alzheimer’s disease identifies new risk loci and implicates Aβ, tau, immunity and lipid processing. Nature Genetics. 2019/03/01 2019;51(3):414-430. doi:10.1038/s41588-019-0358-2

5. Wightman DP, Jansen IE, Savage JE, et al. A genome-wide association study with 1,126,563 individuals identifies new risk loci for Alzheimer’s disease. Nature Genetics. 2021/09/01 2021;53(9):1276-1282. doi:10.1038/s41588-021-00921-z

6. Karlsson IK, Escott-Price V, Gatz M, et al. Measuring heritable contributions to Alzheimer’s disease: polygenic risk score analysis with twins. Brain Commun. 2022;4(1):fcab308. doi:10.1093/braincomms/fcab308

7. Sierksma A, Escott-Price V, De Strooper B. Translating genetic risk of Alzheimer’s disease into mechanistic insight and drug targets. Science. 2020;370(6512):61–66. doi:doi:10.1126/science.abb8575

8. Baker E, Leonenko G, Schmidt KM, et al. What does heritability of Alzheimer’s disease represent? PLoS One. 2023;18(4):e0281440. doi:10.1371/journal.pone.0281440

9. Wightman DP, Jansen IE, Savage JE, et al. A genome-wide association study with 1,126,563 individuals identifies new risk loci for Alzheimer’s disease. Nat Genet. Sep 2021;53(9):1276–1282. doi:10.1038/s41588-021-00921-z

10. Meyer JM, Breitner JCS. Multiple threshold model for the onset of alzheimer’s disease in the NAS-NRC twin panel. American Journal of Medical Genetics. 1998/02/07 1998;81(1):92–97. 10.1002/(SICI)1096-8628(19980207)81:1<92::AID-AJMG16>3.0.CO;2-R

11. Ridge PG, Hoyt KB, Boehme K, et al. Assessment of the genetic variance of late-onset Alzheimer’s disease. Neurobiology of Aging. 2016/05/01/ 2016;41:200.e13-200.e20. 10.1016/j.neurobiolaging.2016.02.024

12. Liu S, Bush WS, Akinyemi RO, et al. Alzheimer disease is (sometimes) highly heritable: Drivers of variation in heritability estimates for binary traits, a systematic review. PLoS Genet. Sep 2025;21(9):e1011701. doi:10.1371/journal.pgen.1011701

13. Richardson K, Norgate S. The equal environments assumption of classical twin studies may not hold. Br J Educ Psychol. Sep 2005;75(Pt 3):339–50. doi:10.1348/000709904x24690

14. Crowley WK. Old Order Amish settlement: diffusion and growth. Annals of the Association of American Geographers. 1978;68(2):249–264.

15. Strauss KA, Puffenberger EG. Genetics, medicine, and the Plain people. Annu Rev Genomics Hum Genet. 2009;10:513–36. doi:10.1146/annurev-genom-082908-150040

16. Lord J, Lu AJ, Cruchaga C. Identification of rare variants in Alzheimer’s disease. Review. Frontiers in Genetics. 2014-October-28 2014;Volume 5 - 2014doi:10.3389/fgene.2014.00369

17. De Deyn L, Sleegers K. The impact of rare genetic variants on Alzheimer disease. Nat Rev Neurol. Mar 2025;21(3):127–139. doi:10.1038/s41582-025-01062-1

18. Steiner MC, Rice DP, Biddanda A, Ianni-Ravn MK, Porras C, Novembre J. Study design and the sampling of deleterious rare variants in biobank-scale datasets. Proceedings of the National Academy of Sciences. 2025;122(23):e2425196122. doi:doi:10.1073/pnas.2425196122

19. Sella G, Barton NH. Thinking About the Evolution of Complex Traits in the Era of Genome-Wide Association Studies. Annual Review of Genomics and Human Genetics. 2019;20(Volume 20, 2019):461-493. 10.1146/annurev-genom-083115-022316

20. Wang Q, Dhindsa RS, Carss K, et al. Rare variant contribution to human disease in 281,104 UK Biobank exomes. Nature. 2021/09/01 2021;597(7877):527–532. doi:10.1038/s41586-021-03855-y

21. Marsh JA, Huang G, Bowling K, et al. Evaluating pathogenicity of variants of unknown significance in APP, PSEN1, and PSEN2. Neurotherapeutics. 2025/04/01/ 2025;22(3):e00527. 10.1016/j.neurot.2025.e00527

22. Cruchaga C, Haller G, Chakraverty S, et al. Rare variants in APP, PSEN1 and PSEN2 increase risk for AD in late-onset Alzheimer’s disease families. PLoS One. 2012;7(2):e31039. doi:10.1371/journal.pone.0031039

23. De Deyn L, Sleegers K. The impact of rare genetic variants on Alzheimer disease. Nature Reviews Neurology. 2025/03/01 2025;21(3):127–139. doi:10.1038/s41582-025-01062-1

24. Guerreiro R, Wojtas A, Bras J, et al. *TREM2* Variants in Alzheimer’s Disease. New England Journal of Medicine. 2013;368(2):117–127. doi:doi:10.1056/NEJMoa1211851

25. Pottier C, Hannequin D, Coutant S, et al. High frequency of potentially pathogenic SORL1 mutations in autosomal dominant early-onset Alzheimer disease. Molecular Psychiatry. 2012/09/01 2012;17(9):875–879. doi:10.1038/mp.2012.15

26. Steinberg S, Stefansson H, Jonsson T, et al. Loss-of-function variants in ABCA7 confer risk of Alzheimer’s disease. Nature Genetics. 2015/05/01 2015;47(5):445-447. doi:10.1038/ng.3246

27. Jonsson T, Atwal JK, Steinberg S, et al. A mutation in APP protects against Alzheimer’s disease and age-related cognitive decline. Nature. 2012/08/01 2012;488(7409):96-99. doi:10.1038/nature11283

28. Bortoluzzi C, Bosse M, Derks MFL, Crooijmans R, Groenen MAM, Megens HJ. The type of bottleneck matters: Insights into the deleterious variation landscape of small managed populations. Evol Appl. Feb 2020;13(2):330–341. doi:10.1111/eva.12872

29. Liu Y, Song YE, Lynn A, et al. Telomere length, aging, and cognitive function in the Midwestern Amish. Human Genetics and Genomics Advances. 2026;7(1)doi:10.1016/j.xhgg.2025.100533

30. Main LR, Song YE, Lynn A, et al. Genetic analysis of cognitive preservation in the midwestern Amish reveals a novel locus on chromosome 2. Alzheimers Dement. Nov 2024;20(11):7453–7464. doi:10.1002/alz.14045

31. Dorfsman D, Prough MB, Gulyayev A, et al. WDR12 and HIVEP3 are contributors to cognitive preservation in Amish SuperAgers. Alzheimer’s & Dementia. 2026/03/01 2026;22(3):e71293. 10.1002/alz.71293

32. Ramos J, Caywood LJ, Prough MB, et al. Genetic variants in the SHISA6 gene are associated with delayed cognitive impairment in two family datasets. Alzheimers Dement. Feb 2023;19(2):611–620. doi:10.1002/alz.12686

33. Taliun D, Harris DN, Kessler MD, et al. Sequencing of 53,831 diverse genomes from the NHLBI TOPMed Program. Nature. 2021/02/01 2021;590(7845):290–299. doi:10.1038/s41586-021-03205-y

34. Das S, Forer L, Schönherr S, et al. Next-generation genotype imputation service and methods. Nature Genetics. 2016/10/01 2016;48(10):1284-1287. doi:10.1038/ng.3656

35. Naj AC, Lin H, Vardarajan BN, et al. Quality control and integration of genotypes from two calling pipelines for whole genome sequence data in the Alzheimer’s disease sequencing project. Genomics. Jul 2019;111(4):808–818. doi:10.1016/j.ygeno.2018.05.004

36. Purcell S, Neale B, Todd-Brown K, et al. PLINK: a tool set for whole-genome association and population-based linkage analyses. Am J Hum Genet. Sep 2007;81(3):559–75. doi:10.1086/519795

37. Manichaikul A, Mychaleckyj JC, Rich SS, Daly K, Sale M, Chen W-M. Robust relationship inference in genome-wide association studies. Bioinformatics. 2010;26(22):2867–2873. doi:10.1093/bioinformatics/btq559

38. Edwards DR, Gilbert JR, Jiang L, et al. Successful aging shows linkage to chromosomes 6, 7, and 14 in the Amish. Ann Hum Genet. Jul 2011;75(4):516–28. doi:10.1111/j.1469-1809.2011.00658.x

39. Teng EL, Chui HC. The Modified Mini-Mental State (3MS) examination. J Clin Psychiatry. Aug 1987;48(8):314–8.

40. Galvin JE, Roe CM, Morris JC. Evaluation of cognitive impairment in older adults: combining brief informant and performance measures. Arch Neurol. May 2007;64(5):718–24. doi:10.1001/archneur.64.5.718

41. Gollan TH, Weissberger GH, Runnqvist E, Montoya RI, Cera CM. Self-ratings of Spoken Language Dominance: A Multi-Lingual Naming Test (MINT) and Preliminary Norms for Young and Aging Spanish-English Bilinguals. Biling (Camb Engl). Jul 2012;15(3):594–615. doi:10.1017/s1366728911000332

42. Morris JC, Heyman A, Mohs RC, et al. The Consortium to Establish a Registry for Alzheimer’s Disease (CERAD). Part I. Clinical and neuropsychological assessment of Alzheimer’s disease. Neurology. 1989;39(9):1159–1165.

43. S.A.G.E. Statistical Analysis for Genetic Epidemiology, Release 6.4.2. 2021;

44. Speed D, Evans DM. Estimating disease heritability from complex pedigrees allowing for ascertainment and covariates. The American Journal of Human Genetics. 2024/04/04/ 2024;111(4):680-690. 10.1016/j.ajhg.2024.02.010

45. Cingolani P, Platts A, Wang le L, et al. A program for annotating and predicting the effects of single nucleotide polymorphisms, SnpEff: SNPs in the genome of Drosophila melanogaster strain w1118; iso-2; iso-3. Fly (Austin). Apr-Jun 2012;6(2):80–92. doi:10.4161/fly.19695

46. Zhou H, Arapoglou T, Li X, et al. FAVOR: functional annotation of variants online resource and annotator for variation across the human genome. Nucleic Acids Res. Jan 6 2023;51(D1):D1300–d1311. doi:10.1093/nar/gkac966

47. Li Z, Li X, Zhou H, et al. A framework for detecting noncoding rare-variant associations of large-scale whole-genome sequencing studies. Nature Methods. 2022/12/01 2022;19(12):1599-1611. doi:10.1038/s41592-022-01640-x

48. STAARpipeline: an all-in-one rare-variant tool for biobank-scale whole-genome sequencing data. Nature Methods. 2022/12/01 2022;19(12):1532-1533. doi:10.1038/s41592-022-01641-w

49. Gogarten SM, Sofer T, Chen H, et al. Genetic association testing using the GENESIS R/Bioconductor package. Bioinformatics. 2019;35(24):5346–5348. doi:10.1093/bioinformatics/btz567

50. Turner SD. qqman: an R package for visualizing GWAS results using Q-Q and manhattan plots. The Journal of Open Source Software. 2018;doi:10.21105/joss.00731

51. Zhou W, Bi W, Zhao Z, et al. SAIGE-GENE+ improves the efficiency and accuracy of set-based rare variant association tests. Nature Genetics. 2022/10/01 2022;54(10):1466-1469. doi:10.1038/s41588-022-01178-w

52. Brandt M. Finding missing heritability in complex traits. Nature Genetics. 2025/12/01 2025;57(12):2942-2942. doi:10.1038/s41588-025-02455-0

53. Wainschtein P, Zhang Y, Schwartzentruber J, et al. Estimation and mapping of the missing heritability of human phenotypes. Nature. 2026/01/01 2026;649(8099):1219-1227. doi:10.1038/s41586-025-09720-6

54. Maaser-Hecker AK, Zellmer JC, Kim M, et al. *RIN3* mutations impairing binding of the Alzheimer&#x2019;s disease-associated protein BIN1 lead to RAB5 hyperactivation and endosomal pathology. Science Advances. 2026;12(5):eadx2127. doi:doi:10.1126/sciadv.adx2127

55. Meshref M, Ghaith HS, Hammad MA, et al. The Role of RIN3 Gene in Alzheimer’s Disease Pathogenesis: a Comprehensive Review. Mol Neurobiol. Jun 2024;61(6):3528–3544. doi:10.1007/s12035-023-03802-0

56. Lopatko Lindman K, Jonsson C, Weidung B, et al. PILRA polymorphism modifies the effect of APOE4 and GM17 on Alzheimer’s disease risk. Scientific Reports. 2022/08/02 2022;12(1):13264. doi:10.1038/s41598-022-17058-6

57. Rathore N, Ramani SR, Pantua H, et al. Paired Immunoglobulin-like Type 2 Receptor Alpha G78R variant alters ligand binding and confers protection to Alzheimer’s disease. PLoS Genet. Nov 2018;14(11):e1007427. doi:10.1371/journal.pgen.1007427

58. Bis JC, Jian X, Kunkle BW, et al. Whole exome sequencing study identifies novel rare and common Alzheimer’s-Associated variants involved in immune response and transcriptional regulation. Molecular Psychiatry. 2020/08/01 2020;25(8):1859-1875. doi:10.1038/s41380-018-0112-7

59. Kunkle BW, Vardarajan BN, Naj AC, et al. Early-Onset Alzheimer Disease and Candidate Risk Genes Involved in Endolysosomal Transport. JAMA Neurology. 2017;74(9):1113–1122. doi:10.1001/jamaneurol.2017.1518

60. Le Guen Y, Belloy ME, Napolioni V, et al. A novel age-informed approach for genetic association analysis in Alzheimer’s disease. Alzheimer’s Research & Therapy. 2021/04/01 2021;13(1):72. doi:10.1186/s13195-021-00808-5

61. Patel T, Brookes KJ, Turton J, et al. Whole-exome sequencing of the BDR cohort: evidence to support the role of the PILRA gene in Alzheimer’s disease. Neuropathol Appl Neurobiol. Aug 2018;44(5):506–521. doi:10.1111/nan.12452

62. Milan KL, Ramkumar KM. Regulatory mechanisms and pathological implications of CYP24A1 in Vitamin D metabolism. Pathology - Research and Practice. 2024/12/01/ 2024;264:155684. 10.1016/j.prp.2024.155684

63. Bivona G, Lo Sasso B, Gambino CM, et al. The Role of Vitamin D as a Biomarker in Alzheimer’s Disease. Brain Sci. Mar 6 2021;11(3)doi:10.3390/brainsci11030334

64. Licher S, de Bruijn RFAG, Wolters FJ, et al. Vitamin D and the Risk of Dementia: The Rotterdam Study. Journal of Alzheimer’s Disease. 2017;60(3):989–997. doi:10.3233/jad-170407

65. Afzal S, Bojesen SE, Nordestgaard BG. Reduced 25-hydroxyvitamin D and risk of Alzheimer’s disease and vascular dementia. Alzheimers Dement. May 2014;10(3):296–302. doi:10.1016/j.jalz.2013.05.1765

66. Feart C, Helmer C, Merle B, et al. Associations of lower vitamin D concentrations with cognitive decline and long-term risk of dementia and Alzheimer’s disease in older adults. Alzheimers Dement. Nov 2017;13(11):1207–1216. doi:10.1016/j.jalz.2017.03.003

67. Alia E, Kerr PE. Vitamin D: Skin, sunshine, and beyond. Clin Dermatol. Sep-Oct 2021;39(5):840–846. doi:10.1016/j.clindermatol.2021.05.025

68. Pandey P, Pradhan S, Mittal B. LRP-associated protein gene (LRPAP1) and susceptibility to degenerative dementia. Genes, Brain and Behavior. 2008/11/01 2008;7(8):943-950. 10.1111/j.1601-183X.2008.00436.x

69. Sánchez L, Alvarez V, González P, González I, Alvarez R, Coto E. Variation in the LRP-associated protein gene (LRPAP1) is associated with late-onset Alzheimer disease. American Journal of Medical Genetics. 2001/01/08 2001;105(1):76-78. 10.1002/1096-8628(20010108)105:1<76::AID-AJMG1066>3.0.CO;2-R

70. Arzouni N, Matloff W, Zhao L, Ning K, Toga AW. Identification of Dysregulated Genes for Late-Onset Alzheimer’s Disease Using Gene Expression Data in Brain. J Alzheimers Dis Parkinsonism. 2020;10(6)

71. Panyard DJ, McKetney J, Deming YK, et al. Large-scale proteome and metabolome analysis of CSF implicates altered glucose and carbon metabolism and succinylcarnitine in Alzheimer’s disease. Alzheimers Dement. Dec 2023;19(12):5447–5470. doi:10.1002/alz.13130

72. Oh S, Jung J, Kim J, et al. Discovery and validation of biomarkers for Parkinson’s disease from human cerebrospinal fluid using mass spectrometry-based proteomics analysis. eBioMedicine. 2025/08/01/ 2025;118:105844. 10.1016/j.ebiom.2025.105844

73. Wang B, Wang XP. Does Ceruloplasmin Defend Against Neurodegenerative Diseases? Curr Neuropharmacol. 2019;17(6):539–549. doi:10.2174/1570159x16666180508113025

74. Liu Y, Song YE, Lynn A, et al. No association of Alzheimer disease with the joint effect of *HFE* and *TF* in the mid-western Amish. Journal of Medical Genetics. 2026;63(2):93. doi:10.1136/jmg-2025-111085

75. Smith MA, Zhu X, Tabaton M, et al. Increased Iron and Free Radical Generation in Preclinical Alzheimer Disease and Mild Cognitive Impairment. Journal of Alzheimer’s Disease. 2010;19(1):363–372. doi:10.3233/jad-2010-1239

