## Supplementary material for "Heritability and rare variant contributions to Alzheimer disease in the Midwestern Amish": All Supplementary

**Table S1. Significant loci observed in GWAS of cognitive preservation (CI vs CU).**

| **CHR** | **POS** | **Variant ID** | **REF** | **ALT** | **Beta** | **p-value** | **Nearest gene** |
| --- | --- | --- | --- | --- | --- | --- | --- |
| 19 | 44885243 | rs283811 | A | G | -0.66 | 1.20 x 10^-9^ | *NECTIN2* |
| 19 | 44887076 | rs283815 | A | G | -0.65 | 1.82 x 10^-9^ | *NECTIN2* |
| 19 | 44891712 | rs184017 | T | G | -0.66 | 1.17 x 10^-9^ | *TOMM40* |
| 19 | 44892457 | rs157581 | T | C | -0.66 | 1.17 x 10^-9^ | *TOMM40* |
| 19 | 44892962 | rs157582 | C | T | -0.66 | 1.17 x 10^-9^ | *TOMM40* |
| 19 | 44893408 | rs59007384 | G | T | -0.66 | 1.17 x 10^-9^ | *TOMM40* |
| 19 | 44908684 | rs429358 | T | C | -0.87 | 2.48 x 10^-12^ | *APOE* |

A negative beta indicates increased cognitive impairment risk.

**Table S2. Significant signals observed in GWAS of AD.**

| **CHR** | **POS** | **Variant ID** | **REF** | **ALT** | **Beta** | **p-value** | **Nearest gene** |
| --- | --- | --- | --- | --- | --- | --- | --- |
| 19 | 44883210 | rs142042446 | G | GTAA | 1.21 | 8.21 x 10^-9^ | *NECTIN2* |
| 19 | 44884202 | rs12972156 | C | G | 1.21 | 8.21 x 10^-9^ | *NECTIN2* |
| 19 | 44884339 | rs12972970 | G | A | 1.21 | 8.21 x 10^-9^ | *NECTIN2* |
| 19 | 44884873 | rs34342646 | G | A | 1.21 | 8.21 x 10^-9^ | *NECTIN2* |
| 19 | 44885243 | rs283811 | A | G | 1.12 | 5.47 x 10^-13^ | *NECTIN2* |
| 19 | 44887076 | rs283815 | A | G | 1.12 | 5.47 x 10^-13^ | *NECTIN2* |
| 19 | 44891079 | rs71352238 | T | C | 1.21 | 8.21 x 10^-9^ | *TOMM40* |
| 19 | 44891712 | rs184017 | T | G | 1.14 | 2.17 x 10^-13^ | *TOMM40* |
| 19 | 44892362 | rs2075650 | A | G | 1.24 | 3.09 x 10^-9^ | *TOMM40* |
| 19 | 44892457 | rs157581 | T | C | 1.14 | 2.17 x 10^-13^ | *TOMM40* |
| 19 | 44892587 | rs34095326 | G | A | 1.29 | 1.62 x 10^-9^ | *TOMM40* |
| 19 | 44892652 | rs34404554 | C | G | 1.24 | 3.10 x 10^-9^ | *TOMM40* |
| 19 | 44892887 | rs11556505 | C | T | 1.24 | 3.10 x 10^-9^ | *TOMM40* |
| 19 | 44892962 | rs157582 | C | T | 1.14 | 2.17 x 10^-13^ | *TOMM40* |
| 19 | 44893408 | rs59007384 | G | T | 1.14 | 2.17 x 10^-13^ | *TOMM40* |
| 19 | 44906745 | rs769449 | G | A | 1.48 | 4.33 x 10^-12^ | *APOE* |
| 19 | 44908684 | rs429358 | T | C | 1.50 | 1.02 x 10^-18^ | *APOE* |
| 19 | 44912456 | rs10414043 | G | A | 1.45 | 1.50 x 10^-11^ | *APOC1* |
| 19 | 44912678 | rs7256200 | G | T | 1.45 | 1.50 x 10^-11^ | *APOC1* |
| 19 | 44912921 | rs483082 | G | T | 0.91 | 1.42 x 10^-9^ | *APOC1* |
| 19 | 44913484 | rs438811 | C | T | 0.91 | 1.42 x 10^-9^ | *APOC1* |
| 19 | 44915533 | rs5117 | T | C | 0.83 | 2.28 x 10^-8^ | *APOC1* |
| 19 | 45696026 | rs62111729 | G | A | 1.17 | 2.75 x 10^-9^ | *QPCTL* |
| 19 | 46280430 | rs62111812 | G | A | 0.98 | 1.17 x 10^-8^ | *HIF3A* |

A positive beta indicates increased AD risk.

**Table S3. Gene-based rare variant association tests for CU vs AD in AD-related genes.**

| **GENE** | **CHR** | **MAC** | **MAC_CU, n (%)** | **MAC_CI, n (%)** | **No. of RV** | **No. of URV** | **Beta_Burden** | **Model 1 p** | **Model 2 p** | **Model 3 p** |
| --- | --- | --- | --- | --- | --- | --- | --- | --- | --- | --- |
| *ABCA7* | 19 | 106 | 33 (31.13) | 73 (68.87) | 6 | 3 | 0.026 | 0.09 | 0.11 | 0.12 |
| *CASP7* | 10 | 44 | 11 (20.62) | 33 (78.38) | 2 | 0 | 0.005 | 0.63 | 0.79 | 0.82 |
| *SORL1* | 11 | 82 | 19 (21.79) | 63 (78.21) | 3 | 3 | -0.004 | 0.99 | 0.99 | 0.99 |

RV, rare variants; MAC, minor allele counts; No. of RV, number of rare variants; No. of URV, number of ultra-rare variants; Beta_Burden: effect size estimate from the burden component of the SKAT-O test (Model 1). p-values were obtained from gene-based SKAT-O tests.

**Table S4. Single-variant association tests for RVs tested in *RIN3* and *PILRA* associated with cognitive preservation.**

| **GENE** | **Variant rsID** | **CHR** | **POS** | **REF** | **ALT** | **Impact** | **P value** | **Beta** |
| --- | --- | --- | --- | --- | --- | --- | --- | --- |
| *RIN3* | rs150221413 | 14 | 92555895 | G | T | Missense variant | 2.65x10^-4^ | -3.98 |
| *RIN3* | rs149740709 | 14 | 92652218 | G | A | Missense variant | - | - |
| *RIN3* | rs12434929 | 14 | 92652887 | G | C | Missense variant | 0.72 | -0.24 |
| *RIN3* | rs147042536 | 14 | 92676516 | T | C | Missense variant | 0.07 | -1.44 |
| *PILRA* | rs376057079 | 7 | 100373659 | G | C | Initiator codon variant | - | - |
| *PILRA* | rs201649203 | 7 | 100397913 | G | C | Splice donor variant | 0.11 | -1.38 |
| *PILRA* | rs20197335 | 7 | 100399797 | G | A | Missense variant | 0.03 | -1.20 |

p values were obtained from single-variant association tests (model 1).

Variants without reported p-values are ultra-rare variants that cannot be tested individually.

**Table S5. Suggestive signal observed in genome-wide, gene-based noncoding RVATs of coding variants in CI vs CU analysis.**

| **Gene** | **chr** | **category** | **MAC** | **# rare** | **STAAR-O p** |
| --- | --- | --- | --- | --- | --- |
| *KLHDC1* | 14 | Promoter_DHS | 56 | 6 | 5.54 x 10^-6^ |
| *TRAF3IP1* | 2 | enhancer_DHS | 219 | 22 | 6.74 x 10^-6^ |
| *ACER2* | 9 | enhancer_DHS | 119 | 8 | 6.13 x 10^-6^ |
| *LINC00922* | 16 | ncrna | 69 | 3 | 1.69 x 10^-4^ |

Three loci showed suggestive associations after accounting for 17,094 genes across 7 annotation masks (suggestive threshold: p = 1/17094/7 = 8.36 x 10^-6^), and ncrna suggestive threshold: 1/2504=4.00 x 10^-4^

**Table S6. Suggestive signal observed in genome-wide, gene-based noncoding RVATs of coding variants in CU vs AD analysis.**

| **Gene** | **chr** | **category** | **MAC** | **# rare** | **STAAR-O p** |
| --- | --- | --- | --- | --- | --- |
| *TRAF3IP1* | 2 | Enhancer_CAGE | 45 | 4 | 3.89 x 10^-6^ |
| *CD3EAP* | 19 | Enhancer_DHS | 136 | 15 | 4.78 x 10^-6^ |
| *PPP1R13L* | 19 | Promoter_DHS | 79 | 5 | 7.67 x 10^-6^ |
| *ELOC* | 8 | UTR | 15 | 4 | 7.87 x 10^-6^ |

Three loci showed suggestive associations after accounting for 17,033 genes across 7 annotation masks (suggestive threshold: p = 1/17033/7 = 8.39 x 10^-6^).

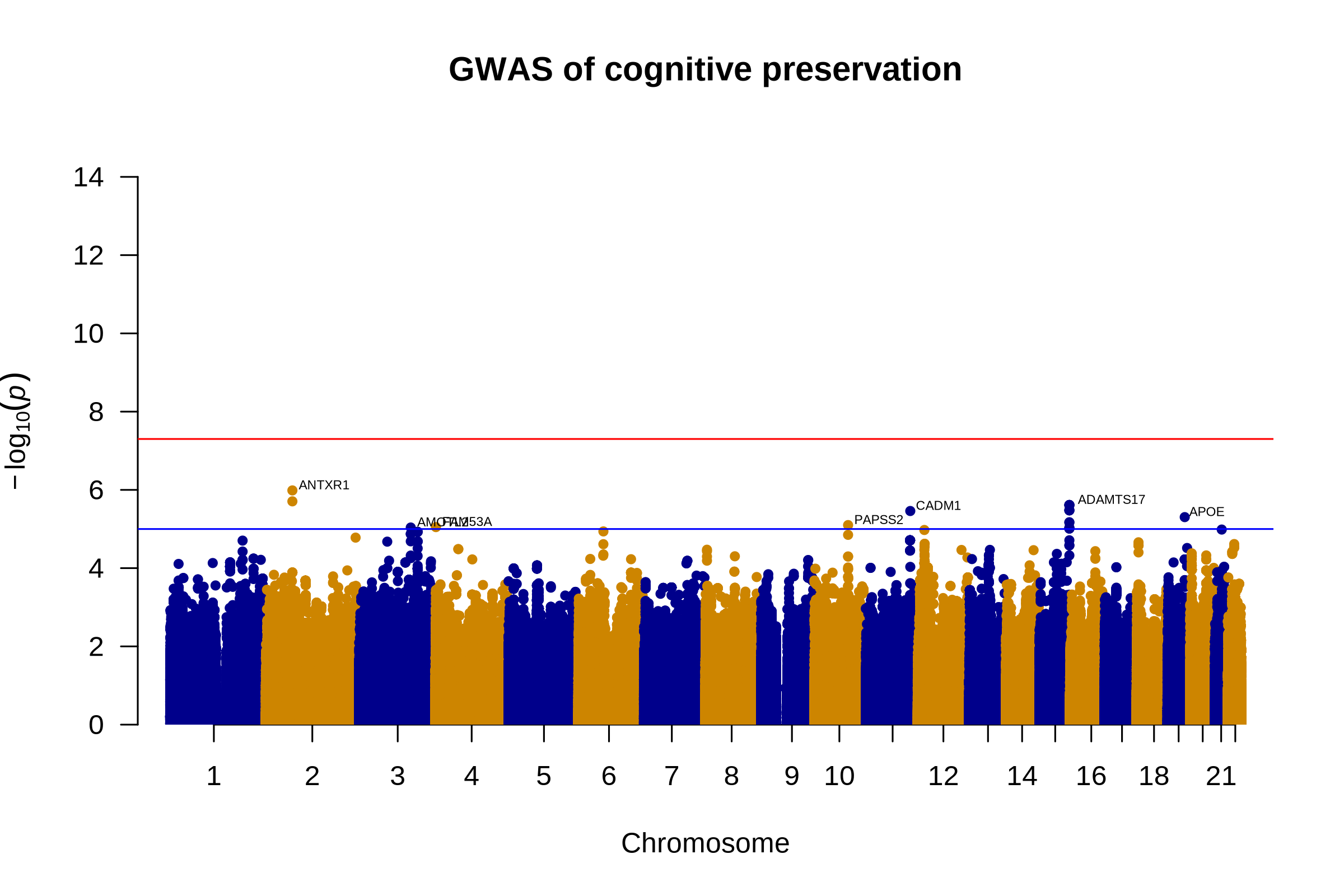

**Figure S1. AD GWAS among APOE ε4 carriers.** Red line: genome-wide significance threshold (−log₁₀ (5 × 10⁻⁸)); blue line: suggestive threshold (−log₁₀(1 × 10⁻⁵)).

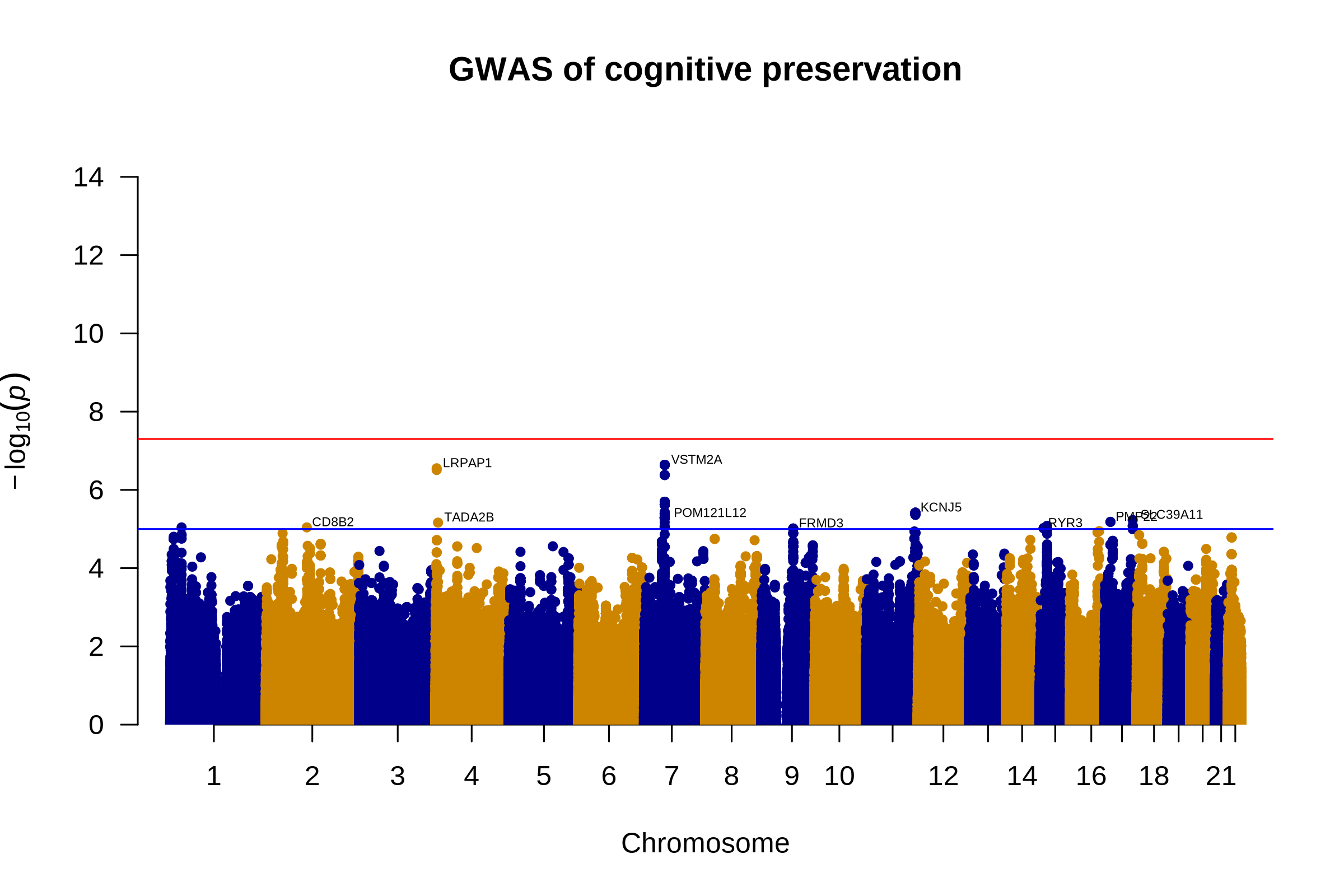

**Figure S2. AD GWAS among non-APOE ε4 carriers.** Red line: genome-wide significance threshold (−log₁₀ (5 × 10⁻⁸)); blue line: suggestive threshold (−log₁₀(1 × 10⁻⁵)).

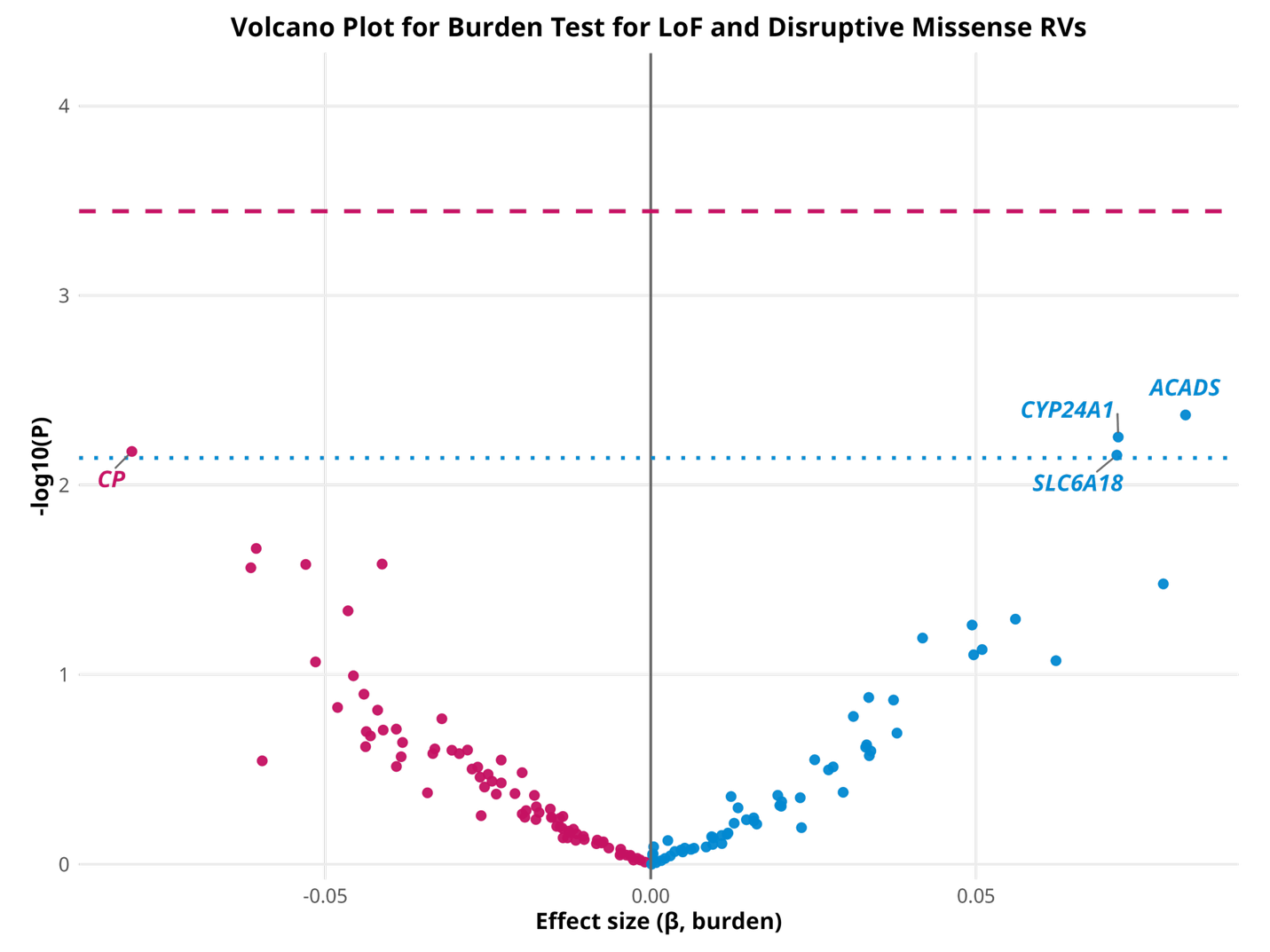

**Figure S3. Burden test of pLoF and disruptive missense for AD among non-APOE ε4 carriers.** 140 genes were tested. Dashed red line: genome-wide significance (−log₁₀(3.57x10^-4^)); dotted blue line: suggestive threshold (−log₁₀(0.00714)). Positive beta indicates increased AD risk.
